# Risk of reinfection after seroconversion to SARS-CoV-2: A population-based propensity-score matched cohort study

**DOI:** 10.1101/2021.03.19.21253889

**Authors:** Antonio Leidi, Flora Koegler, Roxane Dumont, Richard Dubos, María-Eugenia Zaballa, Giovanni Piumatti, Matteo Coen, Amandine Berner, Pauline Darbellay Farhoumand, Pauline Vetter, Nicolas Vuilleumier, Laurent Kaiser, Delphine Courvoisier, Andrew S Azman, Idris Guessous, Silvia Stringhini, SEROCoV-POP study group

## Abstract

**Importance:** Serological assays detecting specific IgG antibodies generated against the Spike protein following Severe Acute Respiratory Syndrome Coronavirus 2 (SARS-CoV-2) infection are being widely deployed in research studies and clinical practice. However, the duration and the effectiveness of the protection conferred by the immune response against future infection remains to be assessed in a large population.

**Objective:** To estimate the incidence of newly acquired SARS-CoV-2 infections in seropositive individuals from a population-based sample as compared to seronegative controls.

**Design:** Retrospective longitudinal propensity-score matched cohort study.

**Setting:** A seroprevalence survey including a population-based representative sample of the population from the canton of Geneva (Switzerland) was conducted between April and June 2020, immediately after the first pandemic wave. Each individual included in the seroprevalence survey was linked to a state centralized registry compiling virologically confirmed SARS-CoV-2 infections since the beginning of the pandemic.

**Participants:** Participants aged twelve years old and over, who developed anti-spike IgG antibodies were matched one-to-two to seronegative controls, using a propensity-score including age, gender, immunodeficiency, body mass index, smoking status and education level.

**Exposure:** SARS-CoV-2 seropositivity.

**Main outcomes and measures:** Our primary outcome was virologically confirmed SARS-CoV-2 infections which occurred from serological status assessment in April-June 2020 to the end of the second pandemic wave (January 2021). Additionally, incidence of infections, rate of testing and proportion of positive tests were analysed.

**Results:** Among 8344 serosurvey participants, 498 seropositive individuals were selected and matched with 996 seronegative controls. After a mean follow-up of 35.6 (Standard Deviation, SD: 3.2) weeks, 7 out of 498 (1.4%) seropositive subjects had a positive SARS-CoV-2 test, of which 5 (1.0%) were considered as reinfections. By contrast, infection rate was significantly higher in seronegative individuals (15.5%, 154/996) during a similar mean follow-up of 34.7 (SD 3.2) weeks, corresponding to a 94% (95%CI 86% to 98%, *P*<0.001) reduction in the hazard of having a positive SARS-CoV-2 test for seropositive subjects.

**Conclusions and relevance:** Seroconversion after SARS-CoV-2 infection confers protection to successive viral contamination lasting at least 8 months. These findings could help global health authorities establishing priority for vaccine allocation.

**Key points:** 

**Question:** Do SARS-CoV-2 antibodies confer protection against future infection?

**Findings:** In this retrospective matched cohort study nested in a representative sample of the general population of Geneva, Switzerland, we observed a 94% reduction in the hazard of being infected among participants with antibodies against SARS-CoV-2, when compared to seronegative controls, >8 months after initial serology assessment.

**Meaning:** Seroconversion to SARS-CoV-2 is associated with a large and sustained protection against reinfection.

## Introduction

SARS-CoV-2 infection induces seroconversion detectable in up to 99% of individuals 2 to 4 weeks following infection with the magnitude of antibody response being influenced by the severity of the disease, timing for testing and the diagnostic performance of immunoassay^1-3^. Markers of cellular and humoral immunity have been shown to last at least 6-8 months after infection^4-7^. However, the extent to which and how long these markers are related to protection against future infections need to be better defined. Because of limited testing availability in the early phases of the pandemic and because of asymptomatic infections, seroprevalence surveys gave the best estimate of the infection’s burden in early phases of pandemic, with rates of seropositivity below 10% after the first wave in Switzerland and in the US^8,9^. Although over 120 million virologically-confirmed SARS-CoV-2 infections have been registered worldwide to date (March 2021), reports of reinfection are scarce and often limited to mild cases^10^, suggesting that SARS-CoV-2 infection elicits protective immunity^11^. Few recent studies among healthcare workers^12-14^, national registries^15-17^ and laboratory de-identified datasets^18^ reported low incidence of reinfection among seropositive or previously infected participants. However, generalisation of their conclusions to the general population is limited by sampling bias (e.g. selection of healthy young participants), lack of control for confounding factors (e.g. no adjustment for comorbidities) or low incidence of infections during the period of observation. To date, no longitudinal assessment of reinfection upon positive serological status has been conducted in representative population-based samples including seniors and comorbid participants.

As in many countries worldwide, in Geneva, Switzerland, the ongoing pandemic has been characterized by two waves, the first reaching its peak in March 2020, the second in November 2020 (Figure 1). In Geneva, the cumulative incidence of virologically confirmed SARS-CoV-2 infections was one of the highest in Europe, with about 8’600 confirmed cases per 100’000 inhabitants, accounted from the beginning of the pandemic in February 2020 to mid-January 2021^19^. With the second wave having occurred 6 to 8 months after the first, we had the opportunity to explore the risk of reinfections in a setting with high community transmission. Here we describe the results of a retrospective cohort study among a population-based sample in Geneva, Switzerland comparing the risk of virologically-confirmed SARS-CoV-2 infections between those with detectable SARS-CoV-2 antibodies after the first pandemic wave to those without.

**Figure 1.**
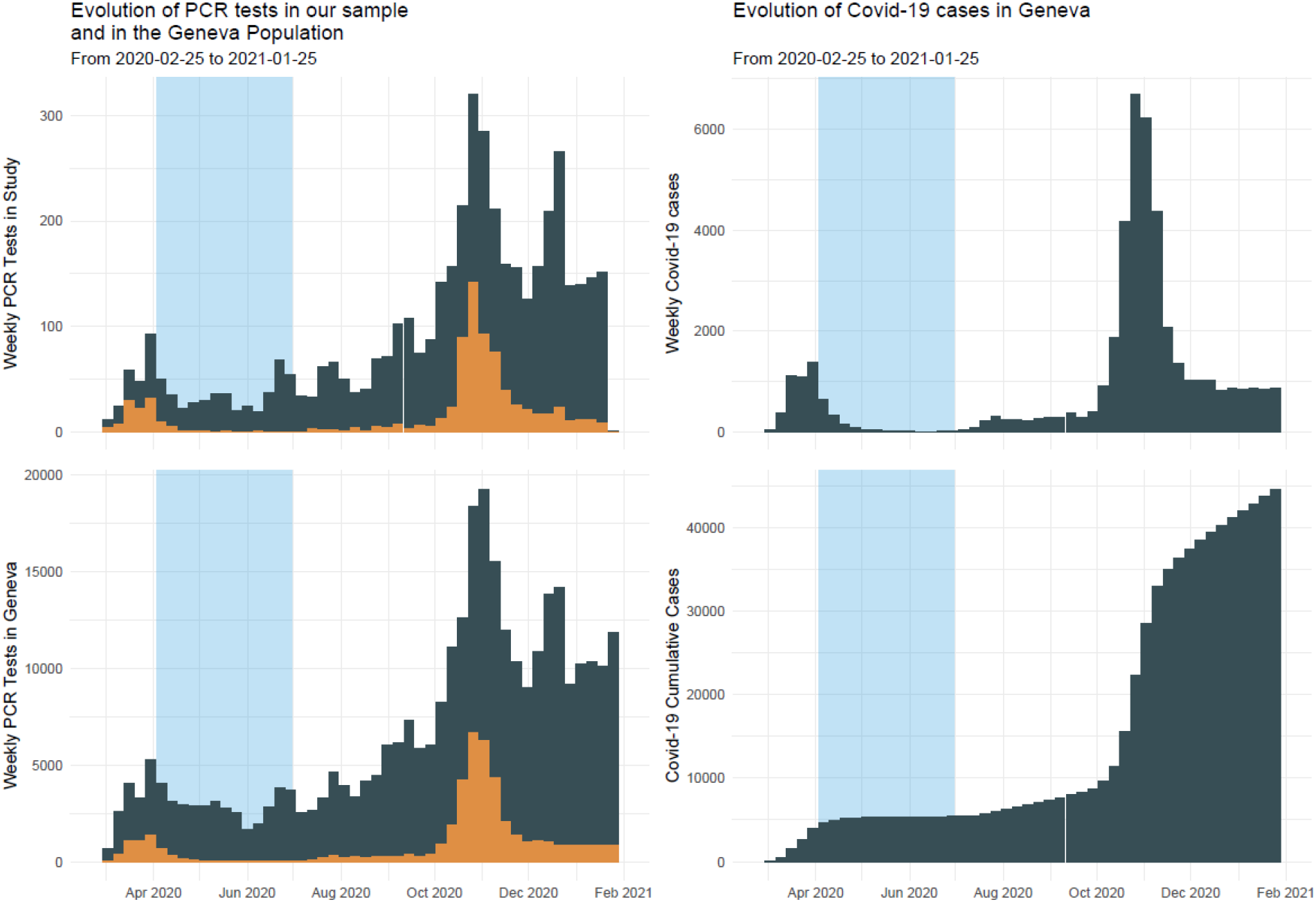
Weekly and cumulative incidence of SARS-CoV-2 infections and incidence of testing during first and second pandemic wave in the canton of Geneva (Switzerland)

**Figure 2.**
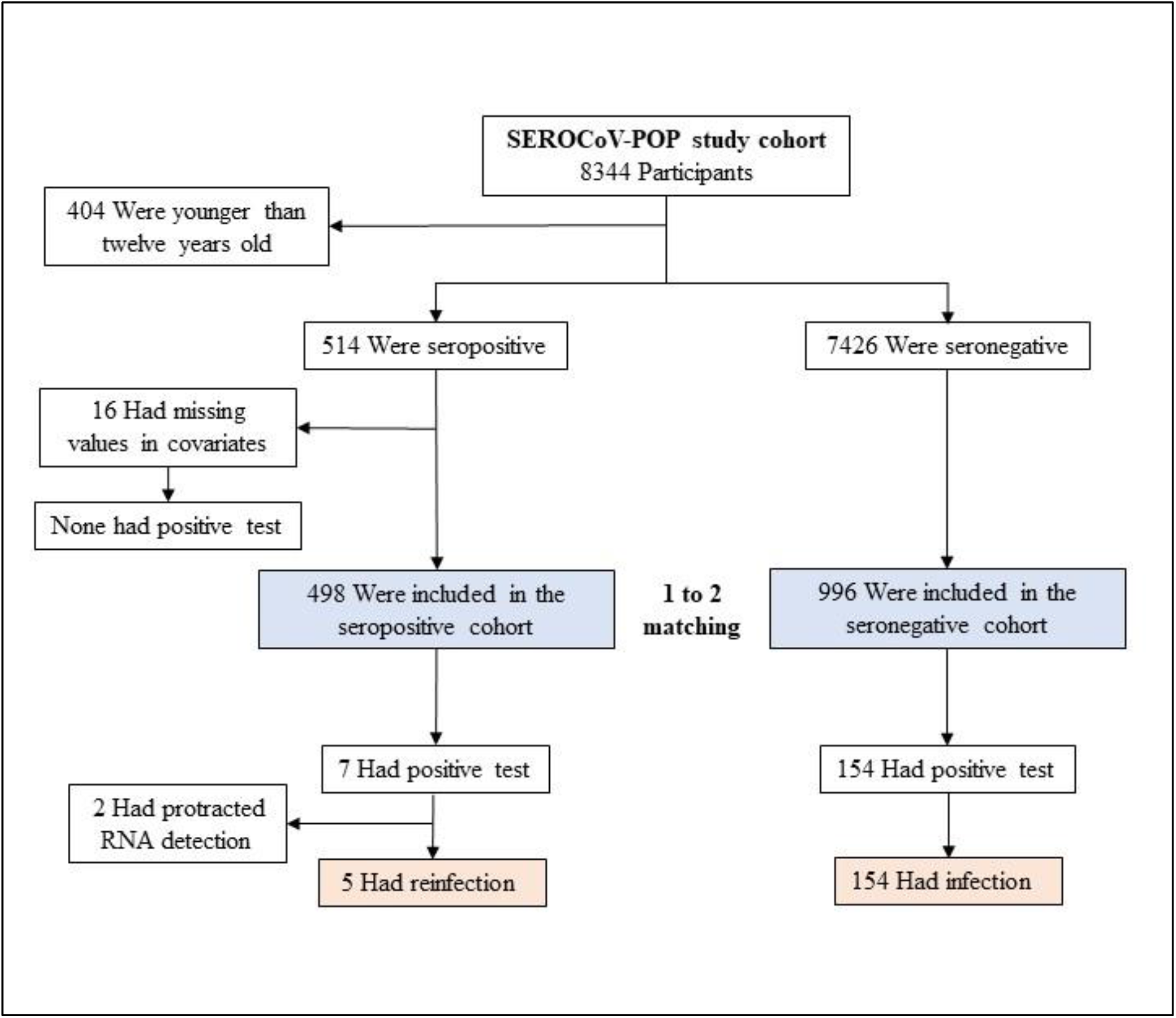
Study flow diagram.

**Figure 3.**
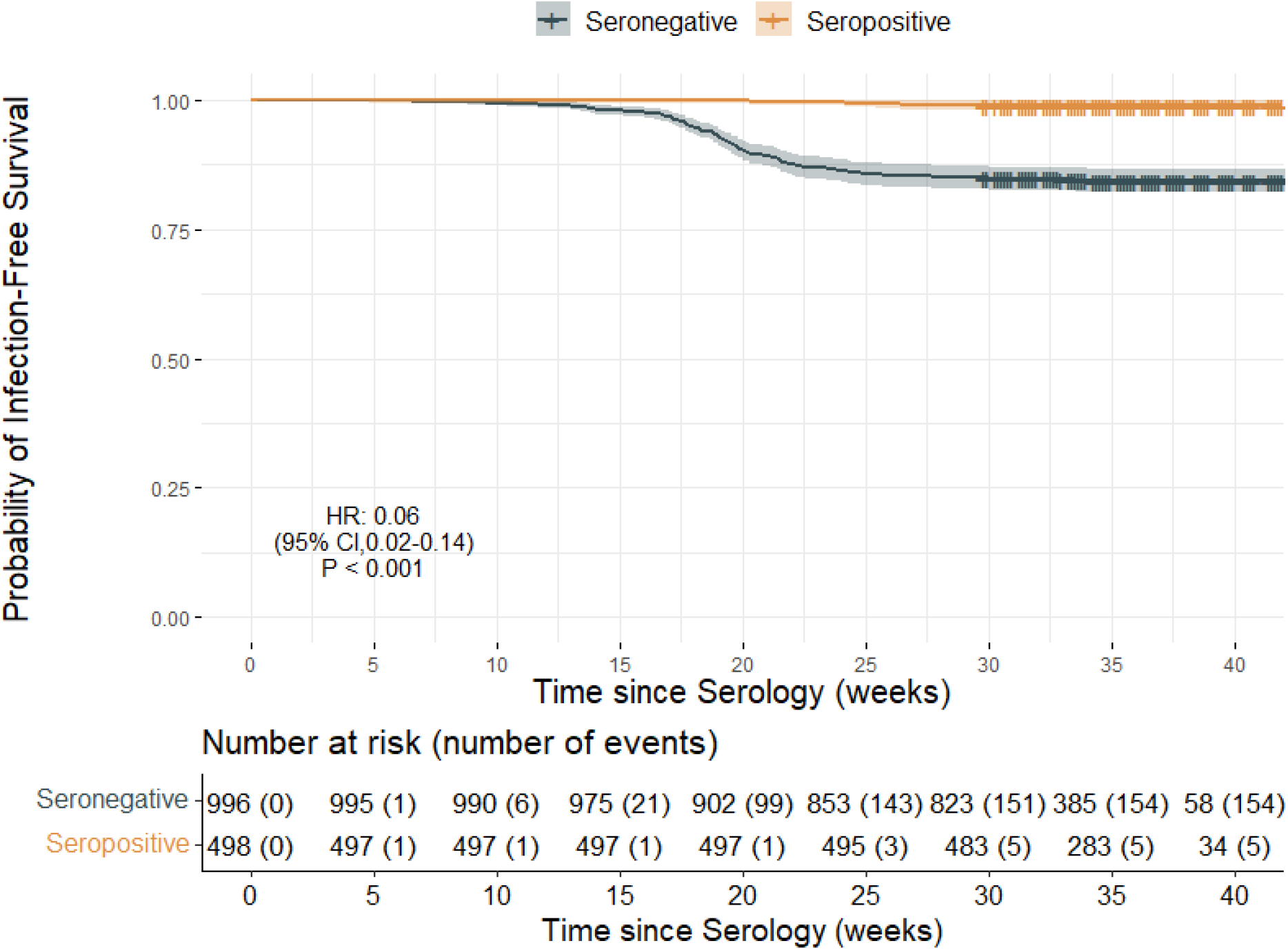
Infection-free survival according to serostatus.

## Methods

### Data sources

#### Seroprevalence survey

From April to June 2020, a representative random sample of adults aged 20 to 74 years old living in the canton of Geneva along with their households was invited to participate in a SARS-CoV-2 serosurvey (SEROCoV-POP). Eight-thousand-three-hundred and forty-four subjects were recruited for an estimated seroprevalence of about 8%; details of sampling and recruitment have been previously described^9,20^.

#### Coronavirus disease 2019 (COVID-19) cases

From the early beginning of the pandemic (February 2020), the General Directorate of Health of Geneva has maintained a centralized registry of SARS-CoV-2 infections and COVID-19 related deaths of the canton. Virologically confirmed SARS-CoV-2 infection was defined by the presence of a positive reverse transcriptase polymerase chain reaction (RT-PCR) or antigenic rapid diagnostic test (Ag RDT) on naso- or oropharyngeal swab, regardless the presence or absence of symptoms.

Communication of testing results and accompanying meta-data was mandatory for all public and private laboratories of the canton. Subjects with multiple testing during the same infective episode (i.e. 90 day time frame after first positive test) were counted as unique cases. During the second pandemic wave, public health authorities recommended the Geneva population to be tested in presence of symptoms or close contact with positive cases and encouraged them with a free from charge policy. As of January 25, 2021, 43’835 cases and 683 deaths had been registered^19^. Data on testing (date and type of test, test result, indication for testing) and COVID-19-related death were extracted and linked on a secured database to each individual included in SEROCoV-POP, using name, sex and date of birth as identity variables for matching.

### Study design and participants

We conducted a retrospective cohort study among subjects 12 years and older enrolled in SEROCoV-POP between April 3 and June 30, 2020. We excluded children 5 to 12 years old as they were unlikely to be tested to SARS-CoV-2, even if symptomatic, based on recommendations from the Swiss Federal Office of Public Health. We classified participants into two groups, according to their baseline antibody status. To limit imbalance in distribution of potential confounders, all seropositive participants were matched to two seronegative controls on baseline characteristics using a propensity score with a logit link function, using a nearest-neighbour distance^21,22^. The propensity score model included variables previously identified as being associated with the risk of developing SARS-CoV-2 infection^23^: age, gender, smoking status (current smoker or e-cigarette user), obesity (body mass index, BMI ≥30 kg/m^2^) and socioeconomic status. The latter was represented in our main analysis by the level of education on a three-level scale: lower (i.e. compulsory education) middle (i.e. secondary education) and higher (i.e. tertiary education). It was replaced by neighbourhood socioeconomic deprivation, as previously described^24^, in one of our sensitivity analyses. Finally, we included self-declared immunodeficiency in the model, as this variable could potentially influence antibodies development, risk of infection and prolonged viral shedding^25^.

### Laboratory analysis

Seropositivity was defined in our primary analysis by the detection of anti-S1 domain of spike protein IgG antibodies using a two-step sequential strategy. Antibodies were first detected by a commercially available ELISA (Euroimmun, Lübeck, Germany #EI 2606-9601 G). All potentially indeterminate (IgG ratio for detection ≥0.5) and positive results were confirmed by a recombinant immunofluorescence assay (rIFA), as this technique was considered the reference method in the laboratory of virology of Geneva University Hospitals (WHO Swiss reference lab) at the time the seroprevalence survey took place. Details on the procedures have been previously described^26^. Additionally, results of ELISA were used independently in our sensitivity analyses, using the manufacturer recommended ratio cut-off for positivity of ≥1.1 (sensitivity 93%, specificity 99%) and an optimized cut-off of ≥2.5 yielding a specificity of 100% against rIFA^26^.

SARS-CoV-2 infection was assessed in the laboratory of virology of Geneva University Hospitals by testing oropharyngeal or nasopharyngeal swabs by RT-PCR using the SARS-CoV-2 reagent kit for BD Max system (Becton, Dickinson and Co, US), the Cobas 6800 SARS CoV2 RT-PCR (Roche, Switzerland), the Xpert Xpress SARS-CoV-2 assay (Cepheid, US) and the TaqPath COVID-19 RT-PCR assay (Thermo Fisher Scientific). Furthermore, results of RT-PCR came from nine private Geneva laboratories licenced by the Swiss Agency for Therapeutic Products, but no details on the diagnostic kits were available. From September 2020 Ag RDT was also implemented as an alternative diagnostic tool in canton of Geneva. Two tests were available: Panbio Covid-19 Ag Rapid Test device (Abbott Diagnostics, Jena, Germany) and Standard Q (SD Biosensor/Roche, Switzerland). Both demonstrated a sensitivity of ≥85% and a specificity of about 100% in an in-house validation study^27^. All SARS-CoV-2 virologic tests (RT-PCR and Ag RDT) were considered equivalent in these analyses.

### Reinfection case classification

Because viral RNA can be detected by RT-PCR more than two months after initial infection^28^, and because swabs collected during the first wave were not available for viral RNA sequencing, all seropositive participants having tested positive for SARS-CoV-2 during follow-up were clinically investigated. Two independent adjudicators with experience in clinical management of SARS-CoV-2 infected patients evaluated suspected cases via hospital electronic health records or phone interview with participants. Adjudication was based on clinical judgement and criteria included, when available, reason for testing, subject’s illness history (including date of symptom onset) and the value and temporal evolution in RT-PCR cycle threshold (Ct). The purpose of this investigation was to differentiate clinical reinfections from protracted RNA detection. Cases of suspected reinfections were classified as *likely* or *unlikely. C*onflicts have been solved by a third person (PDF).

### Outcomes

The primary outcome was a virologically confirmed SARS-CoV-2 infection during the study period, defined by the time between inclusion (date of blood sample collection for serology) and death or the end of the study (January 25, 2021), whichever occurred first. Secondary outcomes were the incidence of virologically confirmed SARS-CoV-2 infections, the test per person ratio and the proportion of positive tests.

### Statistical analysis

We estimated the hazard ratio of having a virologically confirmed SARS-CoV-2 infection comparing those who were anti-S IgG seropositive and seronegative at baseline using survival analysis methods. We used the Kaplan-Meier estimator to estimate the survival functions for each group. We used a Cox’s proportional hazard model with a frailty term for matched set to estimate the hazard ratio. We tested the assumption of proportional hazards using a test based on the Schoenfeld residuals^29^. Protection conferred by seropositivity was estimated computing one minus the hazard ratio. Baseline characteristics, incidence of infections, test/person ratio and test positivity were compared using t-test or McNamar test, as appropriate. A two-sided p value of <0.05 was used to infer statistical significance. A plot of the mean differences between variable was used to evaluate the covariate balance after the matching, with a difference of less than 0.1 considered to be acceptable (eFigure 1 in online-only material).

Consistency of results was assessed for our primary outcome computing hazard ratios for the full unmatched sample, using a univariable and multivariable Cox-proportional hazard model. The same covariates used in the propensity score model were selected for the multivariable model. Additional sensitivity analyses were conducted changing the variables included in the propensity score, varying the definitions of seropositivity, as described above, and considering all suspected cases as reinfections. Statistical analyses were conducted with R, version 4.0.3.

## Ethics

The investigation conforms to the principles outlined in the Declaration of Helsinki and was approved as an amendment by the local Ethical Committee (CCER 2020-00881). Informed consent had been previously obtained from all participants.

## Results

### Study population

From a total of 8344 individuals included in the April-June 2020 serosurvey, 514 aged 12-years-old and older had anti-S1 SARS-CoV-2 IgG antibodies. Sixteen individuals were excluded from the analysis because of missing covariate data. The remaining 498 were matched to 996 seronegative controls (Figure 1). Baseline characteristics were similar between study groups (Table 1). About half of participants were women (51% in both groups), mean age was 46.6 years (SD 16.6) in seropositives and 47.3 years (SD 16.3) in seronegatives. Mean BMI was about 24 kg/m^2^ and distribution of comorbidities was similar between seropositive and seronegative groups, represented mostly by hypertension (9.6% and 9.0% respectively) and chronic pulmonary disease (3.8% and 4.8%). Immunodeficiency was declared by 1.8% of participants in both groups. Most seropositive (91%) and one third of seronegative participants (36.7%) reported at least one COVID-19-related symptom from January 2020 to study inclusion. The list of COVID-19 symptoms is available in Table 1 footnotes.

**Table 1.** Demographics and characteristics of the matched seropositive and seronegative individuals.

|  | Seropositive (n=498) | Seronegative (n=996) | P Value |
| --- | --- | --- | --- |
| **Age, years, mean (SD)** | 46.6 (16.6) | 47.3 (16.3) | - |
| **Age group, years, No. (%)** |  |  |  |
| 12-19 | 32 (6.4) | 61 (6.1) |  |
| 20-39 | 139 (28.0) | 280 (28.1) |  |
| 40-59 | 231 (46.4) | 462 (46.4) |  |
| 60-79 | 90 (18.1) | 181 (18.2) |  |
| ≥ 80 | 6 (1.2) | 12 (1.2) |  |
| **Female, No. (%)** | 256 (51.4) | 510 (51.2) | - |
| **BMI, kg/m<sup>2</sup>, mean (SD)** | 23.8 (4.1) | 23.7 (4.4) | - |
| **Comorbidity, No. (%)** |  |  |  |
| Diabetes | 12 (2.4) | 27 (2.7) | - |
| Hypertension | 48 (9.6) | 96 (9.0) | - |
| Chronic pulmonary disease | 19 (3.8) | 48 (4.8) | - |
| Cancer | 10 (2.0) | 27 (2.7) | - |
| Immunodeficiency | 9 (1.8) | 18 (1.8) | - |
| **Education, No. (%)** |  |  | - |
| Lower education | 40 (8.0) | 77 (7.7) |  |
| Middle education | 190 (38.2) | 381 (38.3) |  |
| Higher education | 268 (53.8) | 538 (54) |  |
| **Follow-up, weeks, mean (SD)** | 35.6 (3.2) | 34.7 (3.2) | <0.001 |
| **Self-reported symptoms before study inclusion, No. (%)** |  |  |  |
| Asymptomatic | 44 (9) | 631 (63.3) | <0.001 |
| Symptomatic<sup>a</sup> | 454 (91) | 365 (36.7) | <0.001 |
| **Rate of testing after study inclusion, mean (SD), No. per individual** | 1.4 (0.7) | 1.5 (0.9) | 0.041 |
| **Proportion of positive test during follow-up, %** | 2.4 % | 11 % | <0.001 |
| **Incidence of virologically confirmed SARS-CoV-2 infections, No. every 1000 person-week (95%CI)** | 0.3 (0.1 to 0.7) | 4.8 (4.6 to 6.2) | <0.001 |

<sup>a</sup> Symptomatic individuals reported at least one COVID-19-related symptom (i.e. fever, cough, cold, throat pain, panting, headache, muscular or articular pain, fatigue, inappetence, nausea, diarrhea, stomach pain, taste and smell loss), from January 2020 to study inclusion. BMI means body mass index,

### Risk of reinfection

After study inclusion, participants were followed-up for a period of 35.6 (SD 3.2) and 34.7 (SD 3.2) weeks in seropositive and seronegative groups, respectively. The testing rate (i.e. test per person ratio) during study follow-up was slightly different between groups, being 1.39 (SD 0.70) and 1.52 (SD 0.87), respectively. Inversely, the proportion of positive test was significantly lower in the seropositive group (2.4% versus 11%, *P* <0.001). The rate of testing was extremely low in both groups before study inclusion, due to restriction in test availability (Figure 1).

Among all seropositive individuals at study inclusion, seven (1.4%) had a subsequent positive RT-PCR testing for SARS-CoV-2. Five of them (1%) were classified as *likely* and two as *unlikely* reinfections by outcome adjudicators (eTable 1 in online-only material) corresponding to an incidence of 0.3 (95%CI 0.1 to 0.7) per 1000 person-weeks. By contrast, the rate of confirmed SARS-CoV-2 infections was significantly higher in seronegative individuals (15.5%, 154/996) corresponding to an incidence rate of 4.8 (95%CI 4.6 to 6.2) per 1000 person-weeks (*P* value for difference <0.001). Over the study follow-up, seropositive individuals were 94% less likely to have a virologically confirmed SARS-CoV-2 infection, when compared to individuals with no detectable anti-SARS-CoV-2 antibodies at study inclusion (hazard ratio of 0.06, 95% CI 0.02 to 0.14, *P*<0.001). Results were consistent in all sensitivity analyses presented in Table 2 and eTable2 (online-only material).

**Table 2.** Number and risk of infection in seropositive and seronegative study groups according to definition of seropositivity.

| Immunoassay strategy | Number of infections, No. (%) |  | Hazard ratio (95%CI) | P value |
| --- | --- | --- | --- | --- |
|  | seropositive | seronegative |  |  |
| Two-step strategy<sup>a</sup> | 5/498 (1.0%) | 154/996 (15.5%) | 0.06 (0.02 to 0.14) | <0.001 |
| EI only, manufacturer cut-off<sup>b</sup> | 12/551 (2.2%) | 200/1102 (18.2%) | 0.11 (0.06 to 0.20) | <0.001 |
| EI only, optimized cut-off<sup>c</sup> | 3/381 (0.8%) | 98/762 (12.8%) | 0.05 (0.01 to 0.14) | <0.001 |

<sup>a</sup> Euroimmun ELISA IgG ratio $\geq 0.5$ and subsequent confirmation with recombinant immunofluorescence assay
<sup>b</sup> Euroimmun ELISA IgG ratio $\geq 1.1$ without confirmation
<sup>c</sup> Euroimmun ELISA IgG ratio $\geq 2.5$ without confirmation

## Discussion

This population-based study including seniors and comorbid participants indicates conclusive evidence that having detectable anti-S1 SARS-CoV-2 antibodies is associated with a 94% significant reduction in the hazard of being tested positive, more than 8 months after initial serologic testing.

Comparable findings have been reported in published reports. In one prospective cohort of UK healthcare workers accounting 1265 seroconverted participants, 2 asymptomatic infections were detected after a 6-months follow-up (incidence rate ratio of 0.12, 95% CI 0.03-0.47)^12^. However, the sample only included healthy working-age participants and the study was conducted over a low incidence period (1.08 positive test every 10’000 days at risk versus 6.8 in our study in seronegative subjects). Similarly, a retrospective analysis of above 130’000 RT-PCR-confirmed SARS-CoV-2 infections in Qatar, observed a rate of reinfection (i.e. positive RT-PCR result ≥45 days after first positive swab) of only 0.05%^15^. Pandemic evolution in this country was monophasic with an intense initial phase affecting mainly young workers and lower incidence of infections from August 2020. The same limitations characterized a subsequent analysis of seroconverted individuals within the Qatar population (only available in preprint form)^16^ limiting generalisation of conclusion to the general population. As the risk of infection is closely tied to intensity of exposure, our study allows to overcome these limitations by sampling a representative sample of the population with a follow-up covering into a high incidence phase having occurred several months after serological assessment.

In a retrospective analysis of a de-identified data from commercial laboratories including over 3 million US individuals, seropositive individuals were less likely of being tested positive to SARS-CoV-2, starting from 30 days after serological testing^17^, although follow-up was limited (median of <2 months) and results were unadjusted for confounders.

Promising preliminary results have been reported for vaccines using lipid nanoparticle-mRNA technology with protection rate above 90% after two injections after a median follow-up of 2 months^30,31^. In the present study, at least a similar rate of protection was conferred by seroconversion and lasted for a mean period of over 8 months.

This study has some limitations. First, not all individuals included in SEROCoV-POP were randomly selected from the community, because original participants were invited to come with their household members. However, characteristics of participants were previously compared with participants of community-surveys in Geneva and did not differ significantly^9^. Second, it is important to consider that although the cantonal testing registry used for identifying infections is wide, the study is limited in being able to only account for infection that have resulted in a test being performed. The number of positive tests depend on screening strategies enacted by health authorities and access to the tests. This may have caused under-detection, despite testing was broadly deployed during the second wave lowering the undetected/detected ratio from 11.6 during the first wave to 2.7 during the second ^32^. That risk of underdetection may be greater in seropositive individuals assuming that reinfections are less symptomatic. Moreover, excessive under-testing during follow-up could potentially have occurred in seropositive individuals because of awareness of their serology result and may have influenced testing behaviour. However, this is not supported by the similar testing rate we observed in the two study groups and the low percentage of positivity in the seropositive group. Third, due to their low incidence during the study period, we cannot infer protection against new SARS-CoV-2 variants^33,34^. In fact, by the end of follow-up, variants with a potential of immune escape were only anecdotally detected by the laboratory of virology of the Geneva University Hospitals. This limits us from generalizing our results to these variants of concern. Fourth, presence or absence of anti-SARS-CoV-2 antibodies was only assessed at one time point and using qualitative methods. This did not allow us to estimate a correlation between antibody titers and risk of reinfection nor determine the proportion of individuals with persistent detectable antibodies at follow-up. Finally, immunoassays could be a source of criticism because of their variable diagnostic performance leading to misclassification bias. In our study, accuracy of commercial serological assays was improved by systematic confirmation of results by rIFA or using a higher ELISA cut off in the sensitivity analyses. It is worth noting that comparable results were obtained in sensitivity analyses using less conservative strategies (i.e commercial ELISA without confirmation with manufacturer cut-offs for positivity).

Our study has also several strengths. First, serological status was determined for all individuals in the early phases of the pandemic allowing a uniform and relatively long longitudinal follow-up. Second, results were concordant across all sensitivity analyses raising robustness of our conclusions. Third, even if no viral RNA sequencing was available for comparison, every case of potential reinfection was identified and individually verified by two independent outcome adjudicators acceding to the complete available clinical information. Fourth, in contrast with the previously cited studies, our sample was composed by a large number of participants aged 60 years or more and included comorbid subjects.

In conclusion, documented SARS-CoV-2 reinfections were exceedingly rare, with an incidence of 0.3 infections for every 1000 persons-week, and none were severe.

Seroconversion after symptomatic or asymptomatic SARS-CoV-2 infection seems to be associated with a 10-fold reduction in risk of successive viral contamination, lasting at least eight months. These findings are of utmost importance for global health authorities facing the challenge of efficiently and rapidly deploy a mass vaccination under dosage shortage and at the same time lifting the restrictions in place.

## Data Availability

Our data are accessible to researchers upon reasonable request for data sharing to the corresponding author.

## Author contributions

The idea for the study originally came from AL. AL, FK, MC carried out the literature search. AL, SS, FK, IG and ASA conceptualized and designed the study. GP and DC oversaw database linkage. FK, AB and PDF conducted clinical investigations. RDum, RDub did data analysis. AL, FK, SS, ASA and MEZ wrote the first draft of the manuscript. All authors have read, critically revised and approved the final version of this manuscript.

## Acknowledgements

We would like to thank Sabine Yerly Ferrillo and Manuel Schibler for the clarifications they gave us on laboratory tests; Aglaé Tardin, Camille Genecand and all the team of General Directorate of Health of Geneva for giving us access and details on ARGOS registry; This study would not have been possible without the contribution of the SEROCoV-POP study group (Silvia Stringhini, Idris Guessous, Andrew S. Azman, Hélène Baysson, Prune Collombet, David De Ridder, Paola d’Ippolito, Matilde D’asaro-Aglieri Rinella, Yaron Dibner, Nacira El Merjani, Natalie Francioli, Marion Frangville, Kailing Marcus, Chantal Martinez, Natacha Noel, Francesco Pennacchio, Javier Perez-Saez, Dusan Petrovic, Attilio Picazio, Alborz Pishkenari, Giovanni Piumatti, Jane Portier, Caroline Pugin, Barinjaka Rakotomiaramanana, Aude Richard, Lilas Salzmann-Bellard, Stephanie Schrempft, Maria-Eugenia Zaballa, Zoé Waldmann, Ania Wisniak, Alioucha Davidovic, Joséphine Duc, Julie Guérin, Fanny Lombard, Manon Will, Antoine Flahault, Isabelle Arm Vernez, Olivia Keiser, Loan Mattera, Magdalena Schellongova, Laurent Kaiser, Isabella Eckerle, Pierre Lescuyer, Benjamin Meyer, Géraldine Poulain, Nicolas Vuilleumier, Sabine Yerly, François Chappuis, Sylvie Welker, Delphine Courvoisier, Laurent Gétaz, Mayssam Nehme, Febronio Pardo, Guillemette Violot, Samia Hurst, Philippe Matute, Jean-Michel Maugey, Didier Pittet, Arnaud G. L’Huillier, Klara M. Posfay-Barbe, Jean-François Pradeau, Michel Tacchino, Didier Trono). Finally, the authors are grateful to all the participants of the Bus Santé study and their household members.

