## Supplementary Material for "Risk of reinfection after seroconversion to SARS-CoV-2: A population-based propensity-score matched cohort study"

### Online-only material

Antonio Leidi[1*] MD, Flora Koegler[1*] MD, Roxane Dumont[2], Richard Dubos[2], María-Eugenia Zaballa[2] PhD, Giovanni Piumatti[2,6] PhD, Matteo Coen[1] PhD, Amandine Berner[1] MD, Pauline Darbellay Farhoumand[1] MD, Pauline Vetter MD[4], Prof Nicolas Vuilleumier[3] MD, Prof Laurent Kaiser[4] MD, Prof Delphine Courvoisier[5] PhD, Andrew S Azman[2] PhD, Prof Idris Guessous[2*] MD, Prof Silvia Stringhini[2*] PhD, SEROCoV-POP study group

**eFigure 1. Diagnostic plot for the propensity score matching**

**Description of reinfected individuals**

**eTable 1. Demographic, clinical and laboratory characteristics of individuals with suspected SARS-CoV-2 reinfection**

**eTable 2. Sensitivity analyses**

### eFigure 1. Diagnostic plot for the propensity score matching

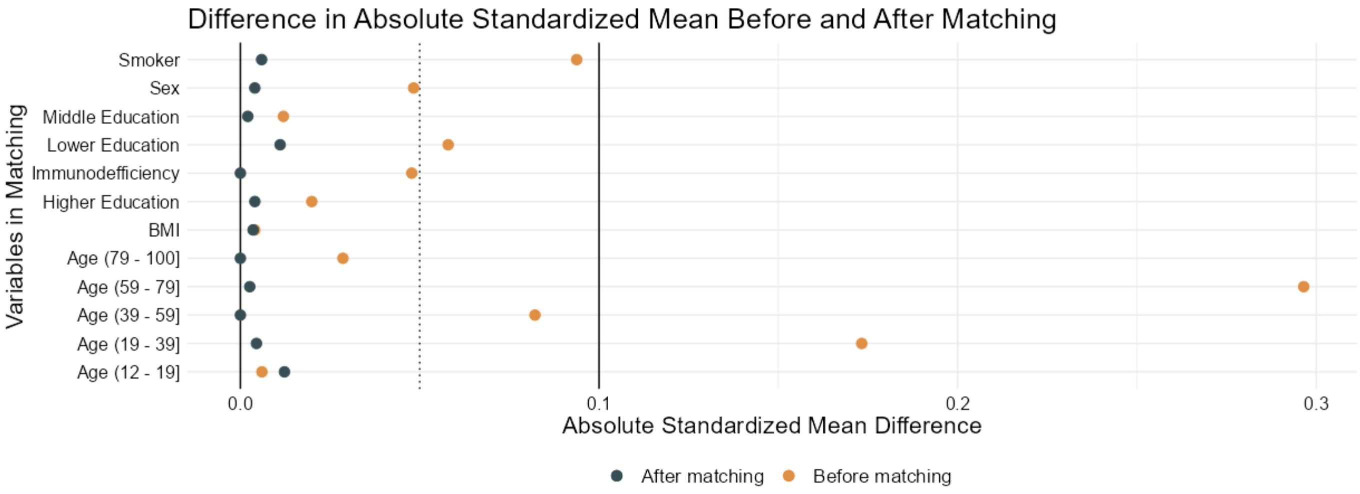


eFigure 1. Standardized mean difference before (orange dots) and after (black dots) propensity score matching. A difference of less than 0.1 considered to be acceptable.

### Description of reinfected individuals

Regarding the five *likely* cases in our main analysis, three of them did not report COVID-like symptoms before serologic testing, whereas reinfections were characterized by classical symptoms. One other seropositive individual was symptomatic at the beginning of the pandemic but was not tested due to restricted testing. She developed light symptoms 120 days after the first infection, with a positive RT-PCR test without informed Ct value. We had no information for the last patient except a 142-day interval between the serologic assay and the positive RT-PCR test. None of these individuals had a RT-PCR testing performed before the serologic assay, and none of them developed a severe form of the disease nor were hospitalized. Regarding the two *unlikely* cases, one individual had a first symptomatic episode confirmed by RT-PCR, without informed Ct value. One-hundred and twenty days later she had a positive RT-PCR test performed for asymptomatic screening, with a Ct value of 34. The other *unlikely* individual had a positive RT-PCR test performed for screening 3 days after the serologic assay. Characteristics of individuals with suspected reinfection included in the main or the sensitivity analyses are reported eTable1.

### eTable 1. Demographic, clinical and laboratory characteristics of individuals with suspected SARS-CoV-2 reinfection

| Patients | Baseline serologic assay | | No of days between epidoses | No of days between serologic assay and reinfection | Clinical characteristics | Timing of PCT, Ct value | Reinfection?[a] | | |
|---|---|---|---|---|---|---|---|---|---|
| | EI | rIFA | | | | | OA1 | OA2 | OA3 |
| **Patient 1: female 42 yrs old** | 0.84 | Positive | unknown | 185 | 1st episode: asymptomatic 2nd episode: symptomatic (mild, COVID-19 - like symptoms: fever, headache, sore throat, dry cough, fatigue, dysgeusia, anosmia) | 1st episode: not done (asymptomatic) 2nd episode: PCR (Cobas 6800 assay), Ct 17.9 | likely | likely | N/A |
| **Patient 2: female 33 yrs old** | 0.58 | Positive | unknown | 178 | 1st episode: asymptomatic 2nd episode: symptomatic (mild, COVID-19 - like symptoms: headache, dysgeusia, anosmia, dyspnea) | 1st episode: not done (asymptomatic) 2nd episode: PCR, no Ct value | likely | likely | N/A |
| **Patient 3: female 77 yrs old** | 0.87 | Positive | unknown | 169 | 1st episode: asymptomatic 2nd episode: symptomatic (mild, COVID-19 - like symptoms: fever, headache, fatigue) | 1st episode: not done (asymptomatic) 2nd episode: PCR, no Ct value | likely | likely | N/A |
| **Patient 4: female 64 yrs old** | 2.0 | Positive | unknown | 142 | 1st episode: no information 2nd episode: asymptomatic (screening) | no information | likely | likely | N/A |
| **Patient 5: female 23 yrs old** | 2.83 | Positive | 120 | 49 | 1st episode: symptomatic (mild, COVID-19 - like symptoms) 2nd episode: asymptomatic (screening) | 1st episode: PCR, no Ct value 2nd episode: PCR (Cobas 6800), Ct 34 | unlikely | likely | unlikely |
| **Patient 6: male 27 yrs old** | 7.6 | Positive | unknown | 3 | 1st episode: no information 2nd episode: asymptomatic (screening) | no information | unlikely | unlikely | N/A |
| **Patient 7: female 68 yrs old** | 1.42 | Positive | 120 | 34 | 1st episode: symptomatic (mild, COVID-19 - like symptoms: dysgeusia, anosmia, fatigue, strong headaches), positive contage with COVID-19. 2nd episode: symptomatic (light, fatigue) | 1st episode: not done (due to testing restriction) 2nd episode: PCR, no Ct value | likely | likely | N/A |
| **Patient 8: female 50 years old** | 2.42 | Negatve | unknown | 132 | 1st episode: asymptomatic 2nd episode: symptomatic (light, COVID-19 - like symptoms: dysgeusia, anosmia, headache) | 1st episode: not done (asymptomatic) 2nd episode: PCR, no Ct value | likely | likely | N/A |
| **Patient 9: female 42 years old** | 1.28 | Negative | unknown | 103 | 1st episode: asymptomatic 2nd episode: symptomatic (light, COVID-19 - like symptoms: fever, dysgeusia, anosmia, headache) | 1st episode: not done (asymptomatic) 2nd episode: PCR, Ct 18.5 | likely | likely | N/A |
| **Patient 10: male 34 years old** | 10.43 | Negative | unknown | 119 | 1st episode: asymptomatic 2nd episode: symptomatic (light, COVID-19 - like symptoms: dysgeusia, anosmia, headache) | 1st episode: not done (asymptomatic) 2nd episode: PCR, no Ct value | likely | likely | N/A |
| **Patient 11: male 19 years old** | 1.38 | Negative | unknown | 150 | 1st episode: no information 2nd episode: symptomatic (light, COVID-19 - like symptoms: fever, muscle pain, fatigue) | 1st episode: not done (asymptomatic) 2nd episode: PCR, Ct 19 | likely | likely | N/A |
| **Patient 12: male 49 years old** | 1.27 | Negative | unknown | 142 | 1st episode: asymptomatic 2nd episode: symptomatic (light, COVID-19 - like symptoms: fever, sore throat, headache, muscle pain) | 1st episode: not done (asymptomatic) 2nd episode: PCR, Ct 16.6 | likely | likely | N/A |
| **Patient 13: male 38 years old** | 1.6 | Negative | unknown | 160 | 1st episode: asymptomatic 2nd episode: symptomatic (light, COVID-19 - like symptoms: fever, headache, dysgeusia, anosmia) | 1st episode: not done (asymptomatic) 2nd episode: PCR, Ct 17.9 | likely | likely | N/A |

| | | | | | | | | | |
|---|---|---|---|---|---|---|---|---|---|
| **Patient 14: male 29 years old** | 1.19 | Negative | unknown | 151 | 1st episode: asymptomatic<br>2nd episode: symptomatic (light, COVID-19 - like symptoms: rhinitis, headache, dyspnea) | 1st episode: not done (asymptomatic)<br>2nd episode: PCR, Ct 20.6 | likely | likely | N/A |
| **Patient 15: male 16 years old** | 1.62 | Negative | no information | 153 | no information | 1st episode: no information<br>2nd episode: PCR, no Ct value | likely | likely | N/A |

[a]Two adjudicators evaluated independently the reinfection probability of each case. Conflicts have been solved by a third adjudicator.

N/A means not applicable, EI means Euroimmun assay IgG ratio; rIFA means recombinant Immunofluorescence assay

### eTable 2. Sensitivity analyses

| | | | | | | |
|---|---|---|---|---|---|---|
| **Number of individuals and infections** | n=7618, events= 618 reinfections=5 | n= 7618, events= 618 reinfections=5 | n= 996, events = 52 reinfections=5 | n= 1494, events= 159 reinfections=5 | n= 1494, events= 161 reinfections=7 | n=1470 events=165 reinfections=5 |
| **Cox model (no imputation on missing data)** | Unmatched univariable | Unmatched multivariable | Matched univariable (1 to 1 ratio) | Matched univariable (2 to 1 ratio) | Matched univariable (2 to 1 ratio) | Matched 2[a] univariable (2 to 1 ratio) |
| **Hazard ratio (95%CI)** | 0.11 (0.05, 0.26) | 0.10 (0.04, 0.25) | 0.10 (0.04, 0.25) | 0.06 (0.02, 0.14) | 0.08 (0.04, 0.18) | 0.06 (0.02, 0.14) |
| ***P* value** | <0.001 | <0.001 | <0.001 | <0.001 | <0.001 | <0.001 |

[a]Neighbourhood socioeconomic deprivation replace Education level in propensity score matching
